## Supplemental file 1 for "How can instructions and feedback with external focus be shaped to enhance motor learning in children? A systematic review"

### S1 file. Search queries for the individual databases

#### PubMed

(("motor learning"[TIAB] OR "procedural learning"[TIAB] OR "declarative learning"[TIAB]) AND (instruction[TIAB] OR instructions[TIAB] OR "internal focus"[TIAB] OR "external focus"[TIAB] OR "focus of attention"[TIAB] OR feedback[TIAB] OR feedback[Mesh] OR "Knowledge of Results (Psychology)"[Mesh]OR "knowledge of results"[TIAB] OR "knowledge of performance"[TIAB] OR "external feedback"[TIAB] OR "learning strategies"[TIAB] OR "learning strategy"[TIAB] OR analogy[TIAB] OR analogies[TIAB] OR "dual task"[TIAB] OR "observational learning"[TIAB] OR observational[TIAB] OR "motor imagery"[TIAB] OR "errorless learning"[TIAB] OR errorless[TIAB] OR "trial and error"[TIAB] OR "guided discovery"[TIAB] OR "differential learning"[TIAB] OR "action observation"[TIAB] OR "practice conditions"[TIAB] OR "random practice"[TIAB] OR "blocked practice"[TIAB] OR "variable practice"[TIAB] OR "repetitive practice"[TIAB] OR "whole practice"[TIAB] OR "part practice"[TIAB] OR "practice schedule"[TIAB] OR "self-controlled practice"[TIAB]))

#### Web of Science

TS=(("motor learning" OR "procedural learning" OR "declarative learning") AND (instruction OR instructions OR "internal focus" OR "external focus" OR "focus of attention" OR feedback OR "knowledge of results" OR "knowledge of performance" OR "external feedback" OR "learning strategies" OR "learning strategy" OR analogy OR analogies OR "dual task" OR "observational learning" OR observational OR "motor imagery" OR "errorless learning" OR errorless OR "trial and error" OR "guided discovery” OR "differential learning" OR "action observation" OR "practice conditions" OR "random practice" OR "blocked practice" OR "variable practice" OR "repetitive practice" OR "whole practice” OR "part practice" OR "practice schedule" OR "self-controlled practice"))

#### Scopus

TITLE-ABS-KEY ( ( {motor learning} OR {procedural learning} OR {declarative learning} ) AND ( instruction OR instructions OR {internal focus} OR {external focus} OR {focus of attention} OR feedback OR {knowledge of results} OR {knowledge of performance} OR {external feedback} OR {learning strategies} OR {learning strategy} OR analogy OR analogies OR {dual task} OR {observational learning} OR observational OR {motor imagery} OR {errorless learning} OR errorless OR {trial and error} OR {guided discovery} OR {differential learning} OR {action observation} OR {practice conditions} OR {random practice} OR {blocked practice} OR {variable practice} OR {repetitive practice} OR {whole practice} OR {part practice} OR {practice schedule} OR {self-controlled practice} ) )

#### Embase

('motor learning'.ti,ab,kw. or exp motor learning/ or 'declarative learning'.ti,ab,kw. or 'procedural learning'.ti,ab,kw.) and (instruction or instructions or 'internal focus' or 'external focus' or 'focus of attention' or feedback or 'knowledge of results' or knowledge of performance' or 'external feedback' or 'learning strategies' or 'learning strategy' or analogy or analogies or 'dual task' or 'observational learning' or observational or 'motor imagery' or 'errorless learning' or errorless or "trial and error" or 'guided discovery' or 'differential learning' or 'action observation' or 'practice conditions' or 'random practice' or 'blocked practice' or 'variable practice' or 'repetitive practice' or 'practice schedule' or 'self-controlled practice' or 'whole practice' or 'part practice).ti,ab,kw.
