## Supplemental file 2 for "How can instructions and feedback with external focus be shaped to enhance motor learning in children? A systematic review"

### S2 file. Excluded studies that nearly met inclusion criteria

#### Excluded because the instructions or feedback with an external focus were compared to instructions or feedback with an internal focus and/or no instructions or feedback


- Bahmani M, Babak M, Land WM, Howard JT, Diekfuss JA, Abdollahipour R. Children's motor imagery modality dominance modulates the role of attentional focus in motor skill learning. Hum Mov Sci. 2021;75: 102742.
- Brocken JEA, Kal EC, van der Kamp J. Focus of attention in children's motor learning: examining the role of age and working memory. J Mot Behav. 2016;48(6): 527–534.
- Chiviacowsky S, Wulf G, Ávila LTG. An external focus of attention enhances motor learning in children with intellectual disabilities. J Intellect Disabil Res. 2013;57(7): 627–634.
- Chow JY, Koh M, Davids K, Button C, Rein R. Effects of different instructional constraints on task performance and emergence of coordination in children. Eur J Sport Sci. 2014;14(3): 224–232.
- Emanuel M, Jarus T, Bart O. Effect of focus of attention and age on motor acquisition, retention, and transfer: a randomized trial. Phys Ther. 2008;88(2): 251–260.
- Gredin V, Williams AM. The relative effectiveness of various instructional approaches during the performance and learning of motor skills. J Mot Behav. 2016;48(1): 86–97.
- Hadler R, Chiviacowsky S, Wulf G, Schild JFG. Children's learning of tennis skills is facilitated by external focus instructions. Motriz Rev Educ Fis. 2014;20(4): 418–422.
- Krajenbrink H, van Abswoude F, Vermeulen S, van Cappellen S, Steenbergen B. Motor learning and movement automatization in typically developing children: the role of instructions with an external or internal focus of attention. Hum Mov Sci. 2018;60: 183–190.

- Lola AC, Tzetzis G. Analogy versus explicit and implicit learning of a volleyball skill for novices: the effect on motor performance and self-efficacy. J Phys Educ Sport. 2020;20(5): 2478–2486.
- Meier C, Fett J, Gröben B. The influence of analogy instruction and motion rule instruction on the learning process of junior tennis players: qualitative assessment of serve performance. Ger J Exerc Sport Res. 2019;49: 291–303.
- Meier C, Frank C, Gröben B, Schack T. Verbal instructions and motor learning: how analogy and explicit instructions influence the development of mental representations and tennis serve performance. Front Psychol. 2020;11: 2.
- Moran KA, Murphy C, Marshall B. The need and benefit of augmented feedback on service speed in tennis. Med Sci Sports Exerc. 2012;44(4): 754–760.
- Parr R, Button C. End-point focus of attention: learning the “catch” in rowing. Int J Sport Psychol. 2009;40(4): 616–635.
- Perreault ME, French KE. Differences in children’s thinking and learning during attentional focus instruction. Hum Mov Sci. 2016;45: 154–160.
- Perreault ME, French KE. External-Focus Feedback Benefits Free-Throw Learning in Children. Res Q Exerc Sport. 2015;86(4): 422–427.
- Roshandel S, Taheri H, Moghadam A. Effects of different attentional focus on learning a motor skill in children. Biosci Res. 2017;14(2): 380–385.
- Teixeira da Silva MBA, Thofehrn Lessa HMS, Chiviacowsky S. Learning of a classical ballet pirouette. J Danc Med Sci. 2017;21(4): 179–184.
- Saemi E, Porter J, Wulf G, Ghotbi-Varzaneh A, Bakhtiari S. Adopting an external focus of attention facilitates motor learning in children with attention deficit hyperactivity disorder. Kinesiology. 2013;45(2): 179–185.

- Schlapkohl N, Tanja H, Raab M. Effects of instructions on performance outcome and movement patterns for novices and experts in table tennis. Int J Sport Psychol. 2012;43(6): 522–541.
- Tse ACY. Effects of attentional focus on motor learning in children with autism spectrum disorder. Autism. 2017;23(2): 405–412.
- Tse ACY, van Ginneken WF. Children's conscious control propensity moderates the role of attentional focus in motor skill acquisition. Psychol Sport Exerc. 2017;31: 35–39.
- Tse ACY, Fong SSM, Wong TWL, Masters R. Analogy motor learning by young children: a study of rope skipping. Eur J Sport Sci. 2017;17(2): 152–159.
- van Cappellen–van Maldegem SJM, van Abswoude F, Krajenbrink H, Steenbergen B. Motor learning in children with developmental coordination disorder: the role of focus of attention and working memory. Hum Mov Sci. 2018;62: 211–220.
- Widenhoefer TL, Miller TM, Weigand MS, Watkins EA, Almonroeder TG. Training rugby athletes with an external attentional focus promotes more automatic adaptions in landing forces. Sports Biomech. 2019;18(2): 163–173.

#### Excluded because the instructions or feedback were applied with reduced frequency, but with an internal focus

- de Carvalho da Silva L, Pereira-Monfredini CF, Teixeira LA. Improved children's motor learning of the basketball free shooting pattern by associating subjective error estimation and extrinsic feedback. J Sports Sci. 2017;35(18): 1–6.
- Weeks DL, Kordus RN. Relative frequency of knowledge of performance and motor skill learning. Res Q Exerc Sport. 1998;69(3): 224–230.

#### Excluded because the instructions or feedback were applied with self-controlled timing, but with an internal focus

- Goudini R, Ashrafpoornavaee S, Farsi A. The effects of self-controlled and instructor-controlled feedback on motor learning and intrinsic motivation among novice adolescent taekwondo players. Acta Gymnica. 2019;49(1): 33–39.
- Lemos A, Wulf G, Lewthwaite R, Chiviacowsky S. Autonomy support enhances performance expectancies, positive affect, and motor learning. Psychol Sport Exerc. 2017;31: 28–34.

#### Excluded because the instructions or feedback were applied in a visual form, but with an internal focus

- Adams D. The relative effectiveness of three instructional strategies on the learning of an overarm throw for force. Phys Educ. 2001;58(2): 67.
- Potdevin F, Vors O, Huchez A, Lamour M, Davids K, Schnitzler C. How can video feedback be used in physical education to support novice learning in gymnastics? Effects on motor learning, self-assessment and motivation. Phys Educ Sport Pedagog. 2018;23(6): 559–574.
- Pasetto SC, Barreiros JMP, Corrêa UC, Freudenheim AM. Visual and kinaesthetic instructional cues and deaf people’s motor learning. Int J Instr. 2020;14(1): 161–180.
- Puklavec A, Antekolović L, Mikulić P. Acquisition of the long jump skill using varying feedback. Croat J Educ. 2021;23(1): 107–132.

#### Excluded because the feedback with an external focus in the controlled group was also applied with reduced frequency

- Petranek LJ, Bolter ND, Bell K. Attentional focus and feedback frequency among first graders in physical education. J Teach Phys Educ. 2018;38(3): 199–206
